# A Longitudinal Seroepidemiology Study to Evaluate Antibody Response to SARS-CoV-2 Virus and Vaccination in Children in Calgary, Canada from July 2020 to April 2022

**DOI:** 10.1101/2022.11.02.22281665

**Authors:** Emily J. Doucette, Joslyn Gray, Kevin Fonseca, Carmen Charlton, Jamil N. Kanji, Graham Tipples, Susan Kuhn, Jessica Dunn, Payton Sayers, Nicola Symonds, Guosong Wu, Stephen B Freedman, James D. Kellner, the Alberta COVID-19 Childhood Cohort (AB3C) Study Team

## Abstract

**Background:** Measurement of SARS-CoV-2 antibody seropositivity is important to accurately understand exposure to infection and/or vaccination in specific populations.

**Methods:** Children with or without prior SARS-CoV-2 infections, was enrolled in Calgary, Canada in 2020. Venous blood was sampled 4 times from July 2020 to April 2022 for SARS-CoV-2 nucleocapsid and spike antibodies. Demographic and clinical information was obtained including SARS-CoV-2 testing results and vaccination records.

**Results:** 1035 children were enrolled and 88.9% completed all 4 visits; median age 9 years (IQR: 5,13); 519 (50.1%) female; and 815 (78.7%) Caucasian. Before enrollment, 118 (11.4%) had confirmed or probable SARS-CoV-2. By April 2022, 39.5% of previously uninfected participants had a SARS-CoV-2 infection. Nucleocapsid antibody seropositivity declined to 16.4% after more than 200 days after diagnosis. Spike antibodies remained elevated in 93.6% of unvaccinated children after more than 200 days after diagnosis. By April 2022, 408 (95.6%) children 12 years and older had received 2 or more vaccine doses, and 241 (61.6%) 5 to 11 year-old children had received 2 vaccine doses. At that time, all 685 vaccinated children had spike antibodies, compared with 94/176 (53.4%) of unvaccinated children.

**Conclusions:** In our population, after the first peak of Omicron variant infections and introduction of COVID-19 vaccines for children, all vaccinated children had SARS-CoV-2 spike antibodies, in contrast to 53.4% of unvaccinated children. It is not yet known whether a high level of seropositivity at a point in time indicates sustained population-level protection against SARS-CoV-2 transmission or severe COVID-19 outcomes in children.

**Summary:** By April 2022, all vaccinated children with or without acquired SARS-CoV-2 infections had spike antibodies, compared with just over one-half of unvaccinated children. It’s not known whether overall seropositivity level in a population indicates sustained protection against severe COVID-19 outcomes.

## Introduction

Following the emergence of severe acute respiratory syndrome coronavirus 2 (SARS-CoV-2), there have been over 613 million confirmed COVID-19 infections worldwide and 4.2 million in Canada (1). Reverse-transcription polymerase chain reaction (RT-PCR) and rapid antigen assays of respiratory samples have facilitated the diagnosis of symptomatic and asymptomatic COVID-19 infections. While RT-PCR is generally accurate for identifying acute cases of COVID-19 (2), there is a role for serologic testing to identify the presence and duration of antibodies following SARS-CoV-2 infection and/or immunization.

Antibodies produced after COVID-19 infections persist for a variable time (3), with no definite level that specifies a correlate of protection (4) that will prevent reinfection. Readily available tests of antibody response to SARS-CoV-2 target the nucleocapsid protein (N), and the spike protein (S) (5). Nucleocapsid antibody is produced after a SARS-CoV-2 infection and spike antibody is produced after vaccination or infection (5). Initial research focused on evaluating testing methods and antibody responses during acute infection and early post-infection (6-8). Data from surveillance studies have documented that nucleocapsid antibodies persist in adults up to 41 weeks post-infection (3,9,10) however less data exists for children (11-18), especially data which include antibodies against variants. While children rarely experience moderate or severe disease (19), infections are frequent. In Canada children under 19 years comprise 21% of the population and 18% of all confirmed COVID-19 infections have occurred in this age group (20,21)

The primary objective of this study was to measure the serial humoral response to SARS-CoV-2 in children in Calgary, Alberta over a two-year period, including those with and without clinically apparent confirmed or probable COVID-19 infection, and those with and without COVID-19 vaccination. The secondary objective was to evaluate the duration of circulating antibodies against SARS-CoV-2 nucleocapsid and spike antibodies in children.

## Methods

### Study Population

The previously described study methods (22) have been updated. All study participants were under 18 years of age, residing in the Calgary area. Two study groups were enrolled. Electronic consent was obtained from the parents of all participating children (or directly from mature minors). The enrollment target was 1000 children.

Group 1 included children with confirmed or probable COVID-19 infection prior to enrollment, as defined by the provincial department of health (Alberta Health) (23). These children were identified by the provincial health services delivery agency, Alberta Health Services (AHS) and invited to participate. Group 2 included children not diagnosed with COVID-19 infections prior to enrollment, and whose families expressed interest in the study through responding to a study announcement posted on social media. Children in this group either had a prior negative PCR test for SARS-CoV-2 or were never tested.

The study received approval from the University of Calgary Conjoint Health Research Ethics Board (CHREB) (REB20-0480).

### Survey, COVID-19 Diagnostic Test Results, and Vaccination Records

Participants or guardians completed an online survey following each study visit that included questions on demographic features, health history, and behaviors related to public health measures during the COVID-19 pandemic. The survey was adapted from the federal COVID-19 Immunity Task Force (24).

The survey also included questions about COVID-19 testing and vaccination. In addition to self-reporting, the results of all laboratory-conducted COVID-19 PCR tests and vaccines received were confirmed from the AHS centralized laboratory database and vaccine registry. Participants were considered immunized if one or more doses of COVID-19 immunization were received more than 14 days prior to blood sampling.

### Serology – Laboratory Methods

Venous blood was collected and sent to the public health laboratory. Samples were tested with two SARS-CoV-2 IgG assays. First, the Abbott ARCHITECT SARS-CoV-2 nucleocapsid IgG assay was used to detect IgG antibodies against the nucleocapsid (N) protein of SARS-CoV-2. Samples with a sample calibration (S/C) value of ≥1.4 were considered positive based on manufacturer’s recommendations (25). Second, the Abbott ARCHITECT SARS-CoV-2 spike IgG II RUO assay was used to detect IgG antibodies to the receptor binding domain (RBD) of the S1 binding subunit of the spike protein of SARS-CoV-2 and was utilized to measure anti-spike antibodies. Samples with a value of ≥50.0 arbitrary units (AU)/mL were considered positive (26).

### Timing of visits

Four visits were conducted, from July 2020 and every 6 months until April 2022. Visit 1 and Visit 2 occurred prior to SARS-Cov-2 variant B.1.617.2 (Delta) being declared a Variant of Concern (VOC) by the World Health Organization (WHO) in May 2021 (27), and prior to vaccine availability for those under 18. Visit 3 occurred after Delta became the dominant variant in Alberta, and after the implementation of COVID-19 vaccines for children ≥12 years, in May 2021. Visit 4 occurred following the emergence of the highly transmissible Omicron variant (27)], and after vaccine implementation for children 5-11 years, in November 2021.

Some children from Group 2 acquired COVID-19 infections in the intervals between visits, and children from both groups may have been reinfected. After each visit, participants were reallocated to Group 3 or 4 depending on their updated infection status. Group 3 consisted of all children who had a confirmed or probable COVID-19 infection before or during the study, and was subdivided into those with infection prior to enrollment (Group 3a, initially Group 1) and those who acquired it between Visit 1 and Visit 4 (Group 3b, initially in Group 2). Group 4 included children who never developed a PCR and/or rapid antigen test confirmed or probable COVID-19 infection throughout the study (initially in Group 2).

By Visit 3 and Visit 4, some children from both groups had been vaccinated against COVID-19. Thus, the results were further stratified by vaccine-relevant age groups and vaccination status. The age groups included <5 years of age (ineligible for vaccine before Visit 4); 5 – 11 years (vaccine-eligible from at Visit 4); and ≥12 years (vaccine-eligible at Visit 3).

### Data Management and Analysis

Electronic consent forms and surveys were hosted on a University of Calgary licensed Research Electronic Data Capture (REDCap) database (28,29). Serology results were compiled using Microsoft Excel 2016 and analyzed using STATA 16 (30) and GraphPad Prism 9.2.0.

The results were initially tabulated separately for Group 1 and Group 2. For the results from Visit 3 and 4, which took place after COVID-19 vaccines were introduced for children and, for Visit 4, after the initial Omicron wave, the results from all groups were compiled and stratified by vaccination status and new infection/reinfection status. In addition, the duration of positive nucleocapsid and spike antibodies after confirmed COVID-19 infections were measured as number of days since the positive PCR test for SARS-CoV-2 antigen for all children for nucleocapsid antibodies, and for all unvaccinated children for spike antibodies.

## Results

### Participants and sampling

A total of 1035 children were enrolled (1023 who attended Visit 1), including 118 (11.4%) with COVID-19 infections prior to enrollment (Group 1) and 917 with not prior infections (Group 2) (Table 1, Figure 1). The median age of participants at enrollment was 9 years (IQR 5-13); there were 519 (50.1%) female participants and 815 (78.7%) self-reported Caucasian ethnicity (Table 1). Of the participants with prior infections, 97 (82%) had a positive PCR test for SARS-CoV-2, while the remaining 21 (18%) were defined as probable COVID-19 infection with a history of symptoms and exposure to a confirmed case.

**Table 1.** Demographic and clinical characteristics of the study population at Visit 1.

| Demographic and Clinical Features | Group 1<br>Prior PCR-confirmed or Probable COVID-19 (n=118) | Group 2<br>No Prior COVID-19 (n=917) | Total (n=1035) | P value |
| --- | --- | --- | --- | --- |
| Age at enrollment, median (IQR), years | 10 (6-14) | 9 (5-13) | 9 (5-13) |  |
| Age group in years at enrollment, n (%), years |  |  |  | 0.033 |
| <5 years | 23 (19.5%) | 178 (19.5%) | 201 (19.4%) |  |
| 5-11 years | 48 (40.7%) | 417 (45.5%) | 464 (44.8%) |  |
| 12+ years | 47 (39.8%) | 322 (35.1%) | 369 (35.7%) |  |
| Total | 118 (100%) | 917 (100%) | 1035 (100%) |  |
| Gender at birth, n (%) |  |  |  | 0.180 |
| • Female | 54 (45.8%) | 465 (50.7%) | 519 (50.1%) |  |
| • Male | 64 (54.2%) | 452 (49.3%) | 516 (49.9%) |  |
| Total | 118 (100%) | 917 (100%) | 1035 (100%) |  |
| Indigenous status, n (%) |  |  |  | 0.453 |
| • Yes | 3 (2.5%) | 33 (3.6%) | 36 (3.5%) |  |
| • No | 105 (89.0%) | 872 (95.1%) | 977 (94.4%) |  |
| Total | 108 (91.5%) | 905 (98.7%) | 1013 (97.9%) |  |
| Ethnicity, n (%) |  |  |  | 0.000 |
| • White | 58 (49.2%) | 757 (82.6%) | 815 (78.7%) |  |
| • Asian (Chinese, Filipino, Japanese, Korean, South Asian, Southeast Asian, West Asian) | 24 (20.3%) | 19 (2.1%) | 43 (4.2%) |  |
| • Black | 4 (3.4%) | 6 (0.7%) | 10 (1.0%) |  |
| • Mixed | 17 (14.4%) | 105 (11.5%) | 122 (11.8%) |  |
| • Other (Arab, Latin American) | 2 (1.7%) | 13 (1.4%) | 15 (1.4%) |  |
| Total | 110 (93.2%) | 900 (98.1%) | 1005 (97.1%) |  |
| Chronic medical conditions, n (%) |  |  |  | 0.354 |
| • Asthma | 8 (6.8%) | 96 (10.5%) | 104 (10.0%) |  |
| • Immune suppressed | 1 (0.8%) | 16 (1.7%) | 17 (1.6%) |  |
| • Chronic neurologic disorder | 1 (0.8%) | 11 (1.2%) | 12 (1.2%) |  |
| • Other Medical Conditions (cancer, chronic blood disorder, chronic heart disease, chronic lung disease, diabetes, hypertension, liver disease) | 0 (0%) | 14 (1.5%) | 14 (1.4%) |  |
| • None of the above | 89 (75.4%) | 595 (64.9%) | 684 (66.1%) |  |

**Figure 1.**
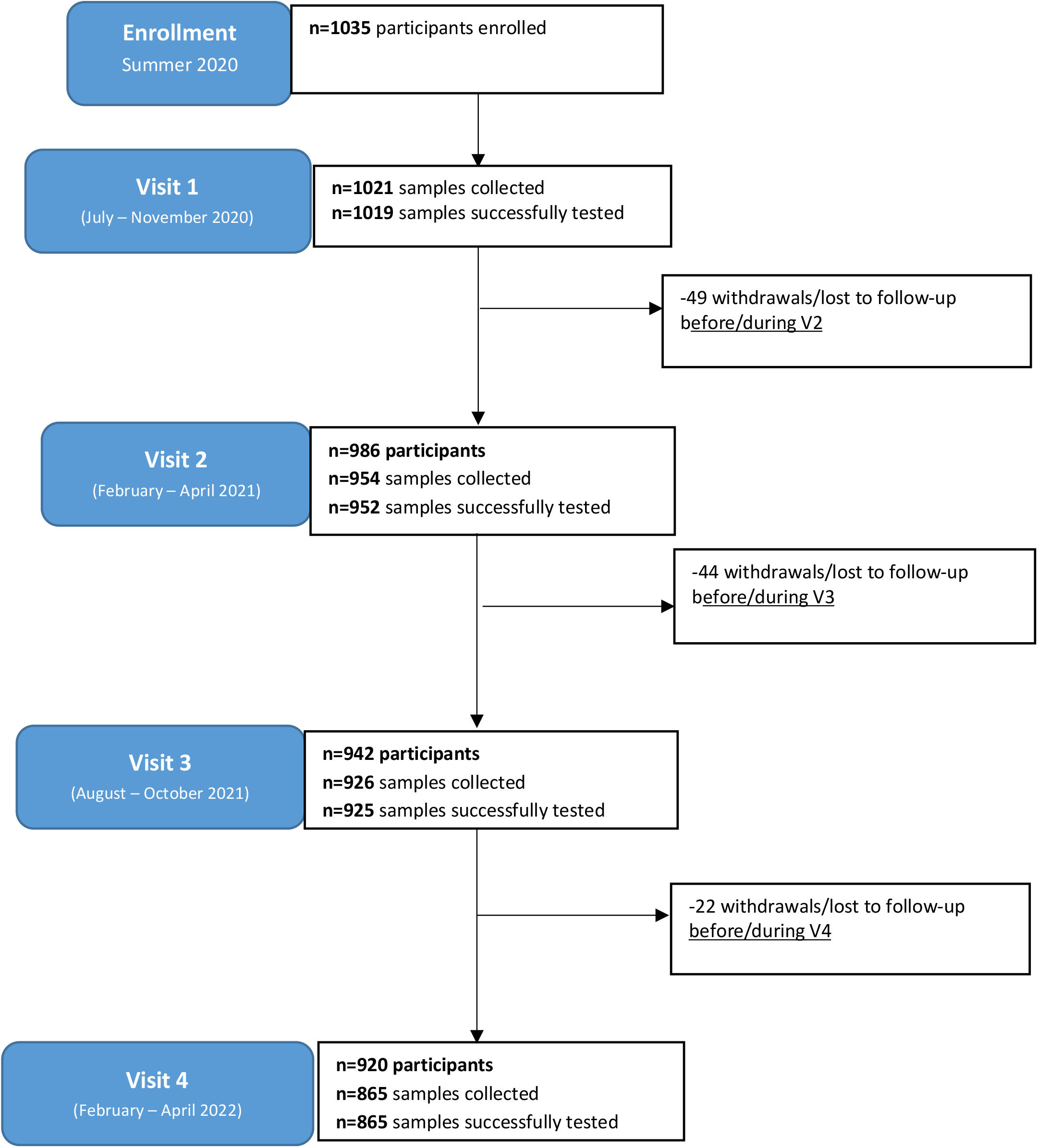
Study participation flow diagram.

There were 3761 blood samples collected over 4 visits. The number of samples successfully tested (and percentage of total initial participants) were: 1019 at Visit 1 (98.5%), 952 at Visit 2 (92.0%), 925 at Visit 3 (89.4%), and 865 at Visit 4 (83.6%) (Figure 1). The number of participants who completed a survey at each visit (and percentage of total participants) were: 994 at Visit 1 (96.0%), 956 at Visit 2 (92.4%), 938 at Visit 3 (90.6%), and 906 at Visit 4 (87.5%). By Visit 4, 115 (11.1%) of the total study population had withdrawn, including 49 at Visit 2, 44 at Visit 3 and 22 at Visit 4. All results prior to withdrawal or loss to follow-up were recorded.

The proportion of all vaccine eligible participants to have received at least one dose of COVID-19 vaccine was 41.8% by visit 3 and 79.2% by visit 4 (Table 2). By Visit 4 which ended in April 2022, the proportion of children immunized was 4.9% (5/102), 79.6% (309/388), and 96.7% (412/426) for children aged < 5 years, 5 – 11 years, and ≥12 years, respectively (Table 2). Although vaccines were not yet approved for children under 5 years, 5 participants were enrolled in an mRNA vaccine clinical trial.

**Table 2.** Vaccine status of participants at Visit 3 and Visit 4 following approval for ages 11+ prior to V3 and ages 5-11 prior to V4 using age at time of visit (vaccination considered effective day 14+ post-immunization).

| Visit 3 (n=942) | Age groups at Visit 3 |  |  |
| --- | --- | --- | --- |
|  | <5 years (n=121) | 5-11 years (n=415) | 12+ years (n=406)<sup>a</sup> |
| Number (%) with 0 doses | 121 (100%) | 394 (94.9%) | 35 (8.6%) |
| Number (%) with 1 dose | 0 | 0 | 19 (4.7%) |
| Number (%) with 2 doses | 0 | 21 (5.1%) | 352 (86.7%) |
| Number (%) with 3 doses | 0 | 0 | 0 |
| Number (%) with unknown vaccine status | 0 | 0 | 0 |
| Visit 4 (n=920) | Age groups at Visit 4 |  |  |
|  | <5 years (n=102) | 5-11 years (n=391)<sup>c</sup> | 12+ years (n=427)<sup>c</sup> |
| Number (%) with 0 doses | 97 (95.1%) | 79 (20.2%) | 14 (3.3%) |
| Number (%) with 1 dose | 1 (0.98%)<sup>b</sup> | 68 (17.4%) | 4 (0.93%) |
| Number (%) with 2 doses | 4 (3.9%)<sup>b</sup> | 241 (61.6%) | 354 (82.9%) |
| Number (%) with 3 doses | 0 | 0 | 54 (12.6%) |
| Number (%) with unknown vaccine status | 0 | 3 (0.8%) | 1 (0.23%) |

<sup>a</sup> Proportion of vaccine eligible participants with $\geq 1$ dose at Visit 3: 41.8%
<sup>b</sup> 5 participants were participating in a COVID-19 vaccine clinical trial for children <5 in parallel to this study.
<sup>c</sup> Proportion of vaccine eligible participants with $\geq 1$ dose at Visit 4: 79.2%

### Incidence of COVID-19 infection during study

Of participants without documented prior COVID-19 infection at enrollment, the number with a newly acquired infection (positive RT-PCR, rapid test, or probable positive), was 0/917 (0%), 15/873 (1.7%), 31/837 (3.7%), and 280/820 (34.1%) at Visits 1, 2, 3 and 4 (Figure 2), respectively. The total cumulative proportion was 39.5% by Visit 4. In addition, of 301 children who reported an infection or reinfection at Visit 4, the proportion diagnosed with only a positive rapid antigen test (185/301; 61.5%) was higher than those with a positive PCR test with or without a rapid test (100/301; 33.2%) or with a probable diagnosis based on exposure to a confirmed case (16/301; 5.3%).

**Figure 2.**
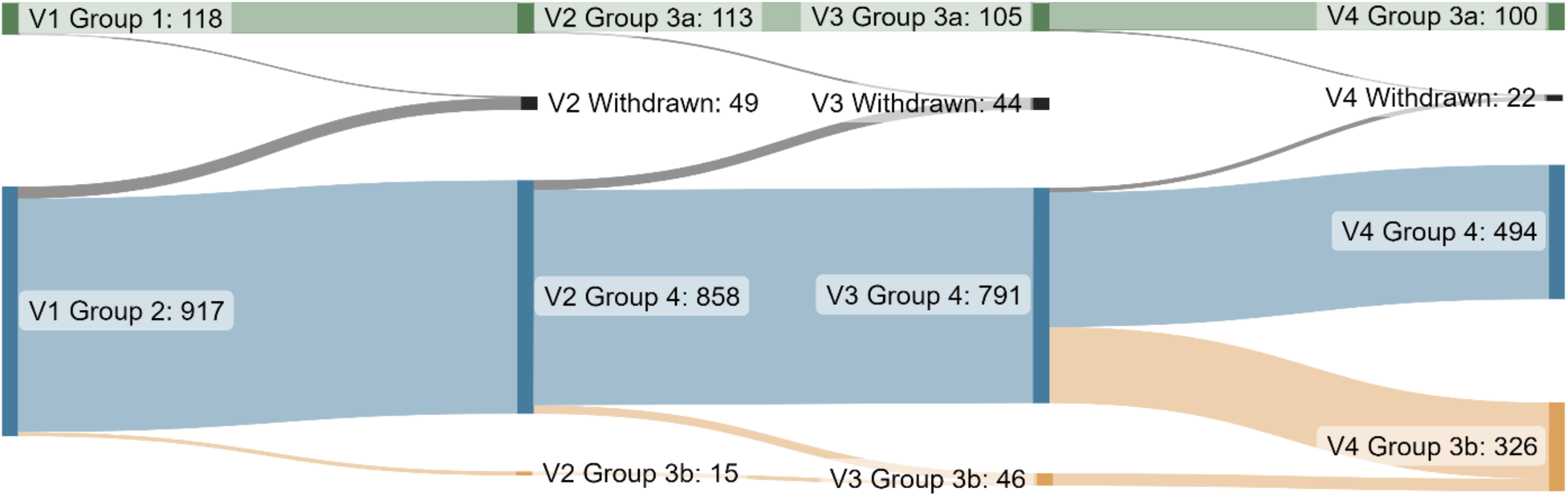
Participant reallocation between groups from Visit 1 (left-hand side) to Visit 4 (right-hand side). Participants with SARS-CoV-2 infection prior to enrollment allocated as having an infection, as shown in Group 1/Group 3a (green). Participants uninfected at enrollment (Group 2) were allocated to Group 4 if they remained uninfected (blue). They were reallocated to Group 3b (orange) if they acquired SARS-CoV-2 infection between study visits. There were 115 total participant withdrawals.

### COVID-19 clinical severity, reinfections, and asymptomatic infections

Study participants reported 1 confirmed hospital admission related to a COVID-19 infection and 3 emergency department visits for symptoms associated with COVID-19 (1 at Visit 1, and 2 at Visit 4). Reinfections (determined by positive PCR or rapid tests) in children with COVID-19 infections prior to enrollment (Group 1) or in children who acquired COVID-19 infections after enrollment (Group 2) were reported for 2 children at Visit 3, and 21 at Visit 4 (18 with prior RT-PCR or rapid test confirmed infections and 3 with RT-PCR or rapid test confirmed infection after a previous probable infection). Neither reinfected participant at Visit 3 was immunized against SARS-CoV-2; however, 66.7% (14/21) of participants by Visit 4 had received at least 1 dose of vaccine.

Asymptomatic infections, defined as newly nucleocapsid antibody-positive participants without a corresponding positive COVID-19 PCR or rapid antigen test (Group 4) were identified in 0.11% (1/909) at Visit 1, 0.84% (7/827) at Visit 2, and 1.9% at Visit 3 (15/779) and 16.8% (77/458) at visit 4. The number of participants with asymptomatic newly positive nucleocapsid antibody who were not immunized prior to sampling was 80.0% (12/15) at Visit 3 and 27.7% (21/77) at Visit 4.

### Seropositivity of Nucleocapsid and Spike antibodies

Table 3 shows the proportions of children with nucleocapsid and spike antibody at each visit stratified by initial allocation to Group 1 (proven or probable COVID-19 prior to enrollment) or Group 2 (no COVID-19 prior to enrollment). From Visits 1 to 3, nucleocapsid antibody seropositivity declined in Group 1 and was low in Group 2, then increased in both groups Visit 4, reflecting the large number of infections or reinfections with the Omicron variant (Table 3, Figure 3). Spike antibody seropositivity remained high in Group 1 throughout the study with a rise at Visit 4 reflecting reinfection and/or vaccination. In Group 2, spike antibody seropositivity increased considerably at Visit 3, reflecting vaccination, and increased more at Visit 4, reflecting vaccination and new infections (Table 3).

**Table 3.**
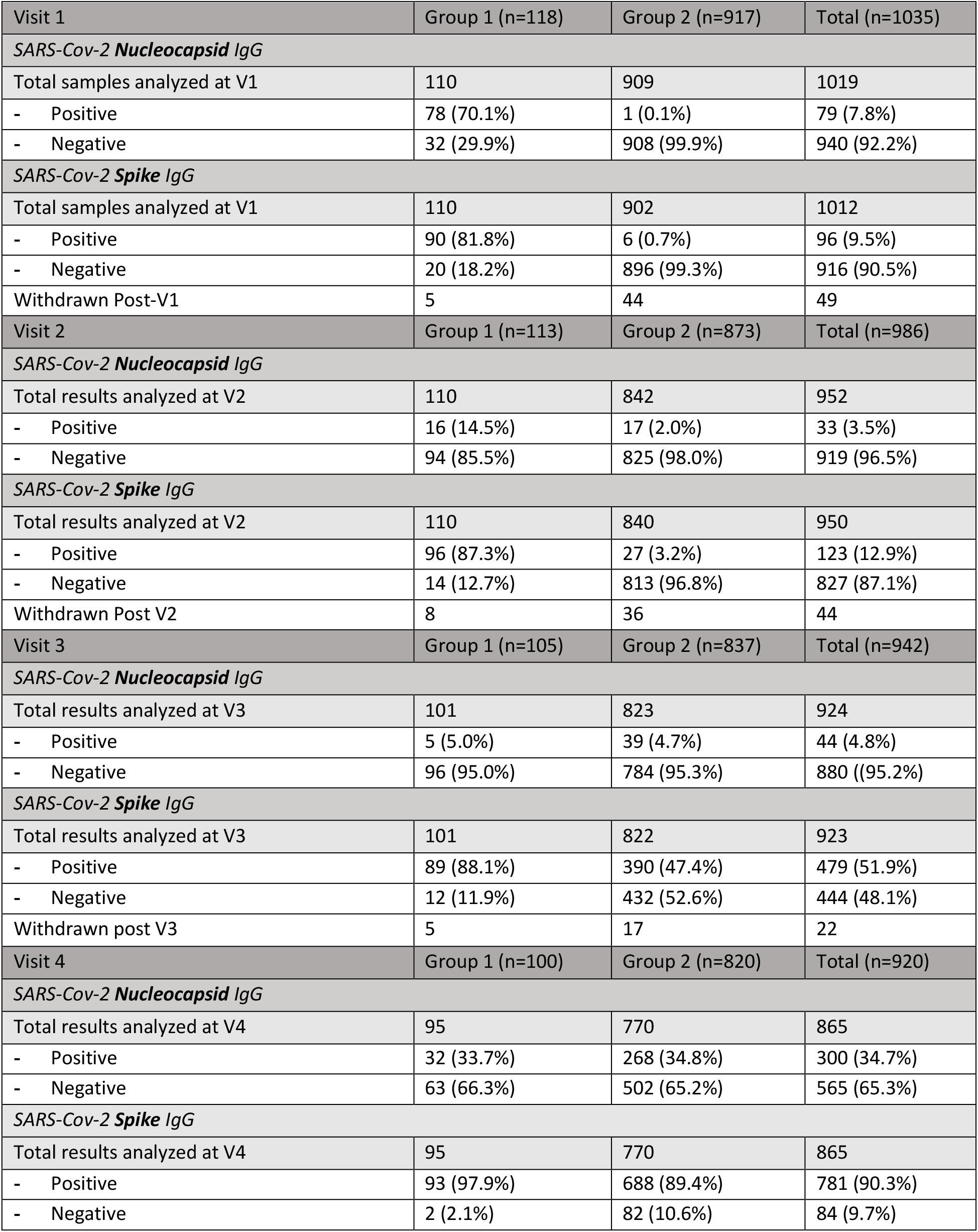
Serology results from Visits 1-4 for initial Group 1 and Group 2.

**Figure 3.**
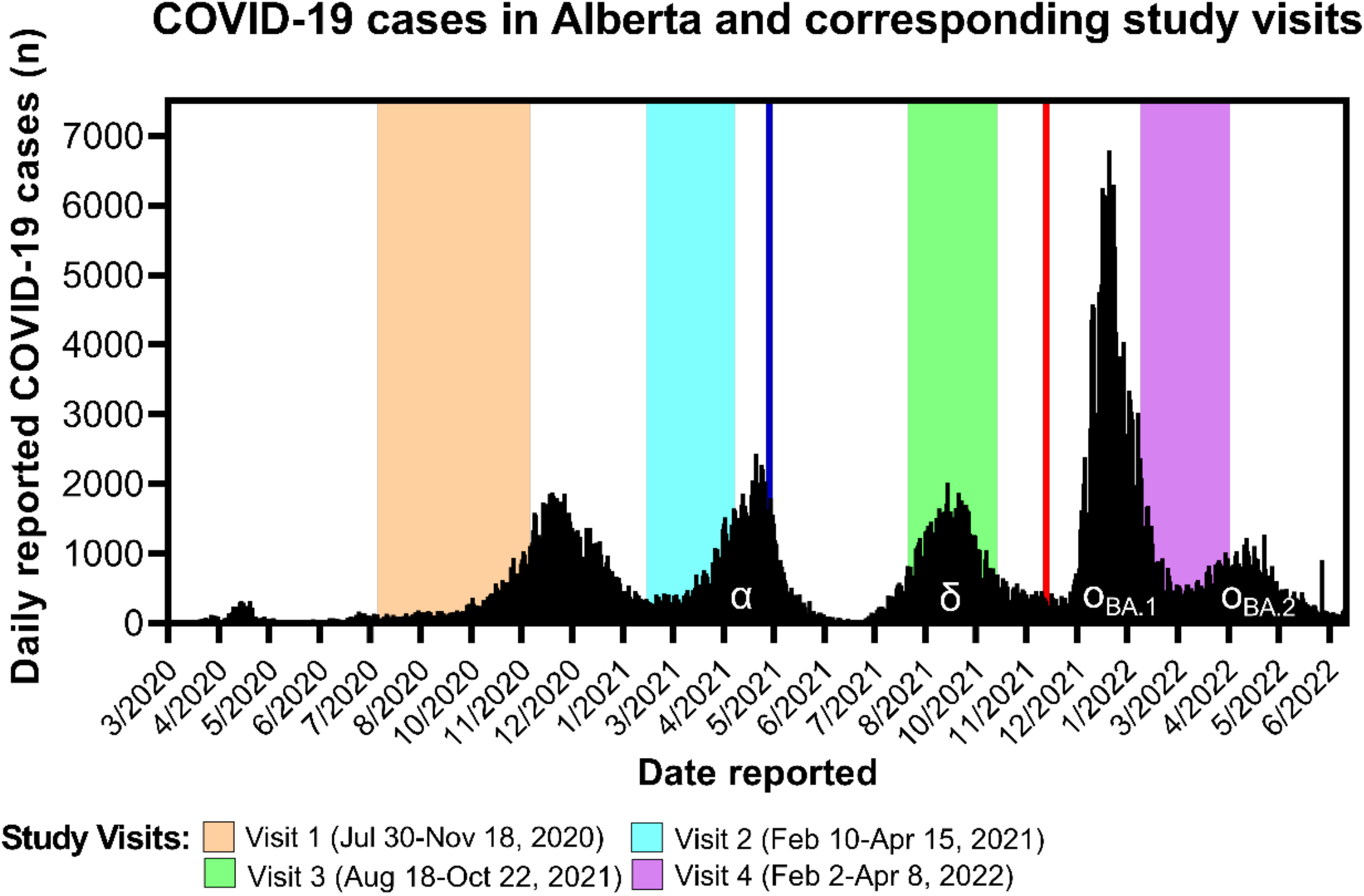
Study visit collection periods with overlay of COVID-19 pandemic curve. Date of vaccine availability for children ≥12 (May 10, 2021) indicated by vertical blue line and for children 5-11 years (November 26, 2021) indicated by vertical red line. Main variant of concern indicated by Greek letter α (Alpha B.1.1.7), δ (Delta B.1.617.2), and ο (Omicron B.1.1.529).

Table 4 shows the increases in nucleocapsid antibody seropositivity at Visit 4 and spike antibody seropositivity samples at Visit 3 and Visit 4 are presented for all participants, stratified by age and vaccination status. At Visit 4, the proportion of all children with positive nucleocapsid antibodies was similar in unvaccinated and vaccinated children (33.5% and 34.9%, respectively). In contrast, 100% of vaccinated children had spike antibodies by Visit 4, compared with 53.4% of unvaccinated children (Table 4).

**Table 4.** Visit 3 and Visit 4 serology results stratified by participant age and vaccine status.

| Visit 3 |  |  |  |  |  |  |  |  |
| --- | --- | --- | --- | --- | --- | --- | --- | --- |
| Age at V3 | All participants, n (%) |  | <5 years, n (%) |  | 5-11 years, n (%) |  | 12+ years, n (%) |  |
| Vaccine Status at V3 | 0 doses | ≥1 dose | 0 doses | ≥1 dose | 0 doses | ≥1 dose | 0 doses | ≥1 dose |
| Total Nucleocapsid Results | 537 | 387 | 120 | N/A | 382 | 21 | 35 | 366 |
| - Positive | 33 (6.1%) | 11 (2.8%) | 12 (10.0%) | N/A | 15 (3.9%) | 0 | 6 (17.1%) | 11 (3.0%) |
| - Negative | 504 (93.9%) | 376 (97.2%) | 108 (90.0%) | N/A | 367 (96.1%) | 21 (100.0%) | 29 (82.9%) | 355 (97.0%) |
| Total Spike Results | 537 | 386 | 119 | N/A | 383 | 21 | 35 | 365 |
| - Positive | 93 (17.3%) | 386 (100.0%) | 22 (18.5%) | N/A | 56 (14.6%) | 21 (100.0%) | 15 (42.9%) | 365 (100.0%) |
| - Negative | 444 (82.7%) | 0 | 97 (81.5%) | N/A | 327 (85.4%) | 0 | 20 (57.1%) | 0 |
| Visit 4 |  |  |  |  |  |  |  |  |
| Age at V4 | All participants, n (%) |  | <5 years, n (%) |  | 5-11 years, n (%) |  | 12+ years, n (%) |  |
| Vaccine status at V4 | 0 doses | ≥1 dose | 0 doses | ≥1 dose | 0 doses | ≥1 dose | 0 doses | ≥1 dose |
| Total Nucleocapsid Results | 176 | 685 | 89 | 5 | 74 | 290 | 13 | 390 |
| - Positive | 59 (33.5%) | 239 (34.9%) | 25 (28.1%) | 1 (20.0%) | 26 (35.1%) | 98 (33.8%) | 8 (61.5%) | 140 (35.9%) |
| - Negative | 117 (66.5%) | 446 (65.1%) | 64 (71.9%) | 4 (80.0%) | 48 (64.9%) | 192 (66.2%) | 5 (38.5%) | 250 (64.1%) |
| Total Spike Results | 176 | 685 | 89 | 5 | 74 | 290 | 13 | 390 |
| - Positive | 94 (53.4%) | 685 (100.0%) | 39 (43.8%) | 5 (100.0%) | 46 (62.2%) | 290 (100.0%) | 9 (69.2%) | 390 (100.0%) |
| - Negative | 82 (46.6%) | 0 | 50 (56.2%) | 0 | 28 (37.8%) | 0 | 4 (30.8%) | 0 |

Note: N/A indicates samples collected during time period when vaccine was not available for children aged 5-11.

### Nucleocapsid and Spike antibody duration

Most, but not all children with RT-PCR confirmed SARS-CoV-2 infections had positive nucleocapsid and spike antibodies at some point in time. More than 150 days after diagnosis, the proportion with nucleocapsid antibodies declined (Figure 4a). In contrast, the proportion of unvaccinated children with spike antibodies did not decline after more than 200 days (Figure 4b) and all 24 children measured from 451 days to more than 700 days after infection remained anti-spike antibody positive.

**Figure 4.**
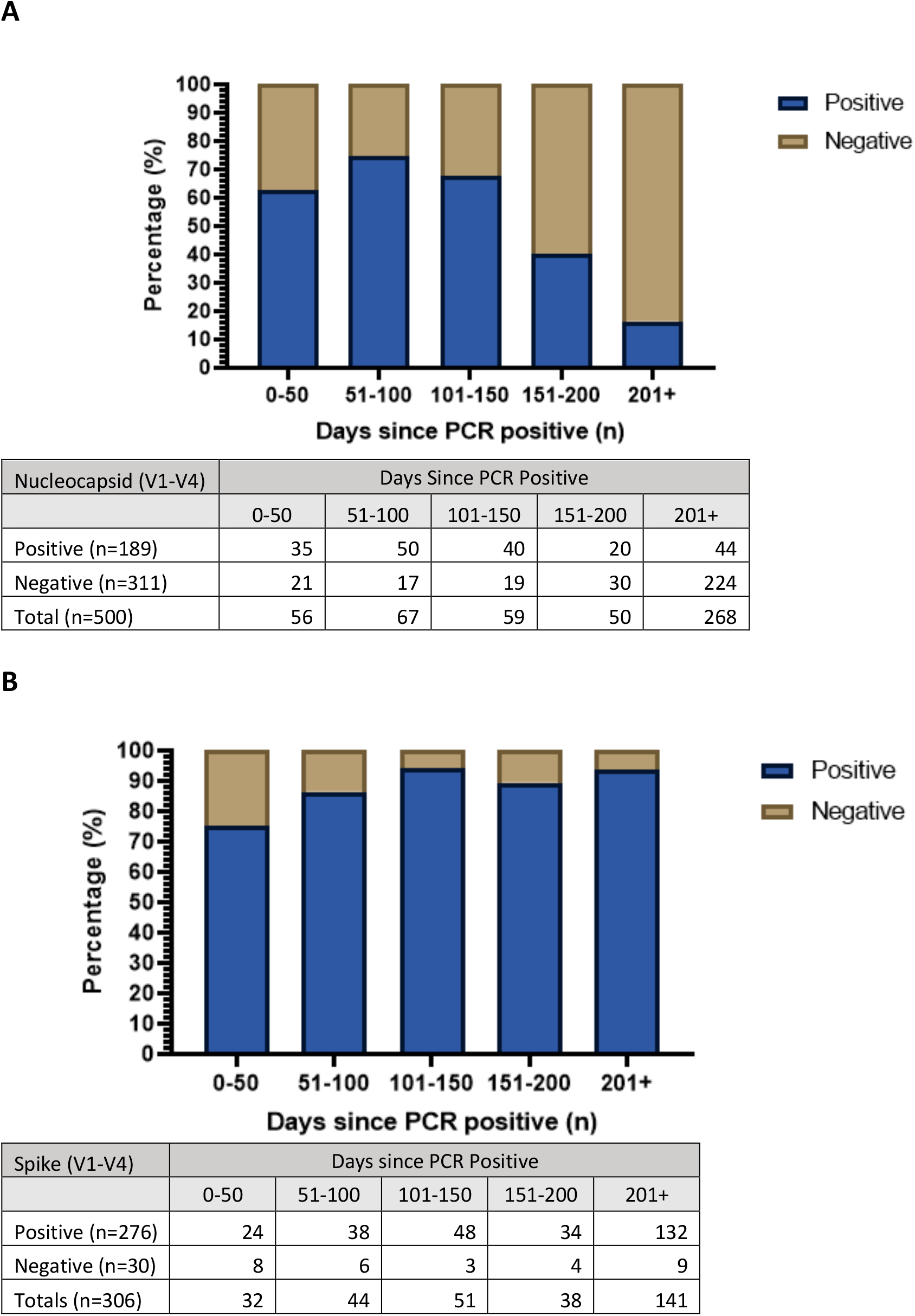
Duration of antibodies in children with confirmed SARS-CoV-2 infection. **(A)** Duration of nucleocapsid antibodies in <u>all children</u> from days since AHS-confirmed PCR positive to subsequent serology test(s), n=500 samples included. (n=19 re-infections and serology results excluded). **(B)** Duration of spike antibodies in <u>unvaccinated children</u> from days since AHS-confirmed PCR positive to subsequent serology test(s), n=306 samples included. (n=19 samples excluded from re-infections, n=61 and n=131 samples excluded at V3 and V4 respectively following vaccination of participants).

## Discussion

We completed SARS-CoV-2 IgG serology testing on over 1000 children on four occasions from July 2020 - April 2022, sampling through successive waves of the pandemic. Before introduction of COVID-19 vaccines for children and youth under 18 years of age, and before the Omicron wave starting in late December 2021, few children seroconverted against SARS-CoV-2 from symptomatic or asymptomatic infection. Seropositivity data on pediatric populations is limited, however our results are consistent with Canadian adult literature from the same period before the Omicron waves (31).

By April 2022, after the start of COVID-19 vaccine programs for children ≥5 years and the first peak of Omicron infections, more than one-third of our study population had serologic evidence of prior infection and 90% had serologic evidence of prior infection and/or immunization. Multiple studies have shown comparable rates of symptomatic infection following Omicron spread (32,33) due to its increased transmissibility and the decreased response from vaccines, although risk for hospitalization may be decreased and milder outcomes are reported (34). It is important to emphasize that the estimate of COVID-19 infection, on the basis of nucleocapsid antibody positivity, is generalizable to the whole population of our province (since Omicron infections occurred in about equal proportions in both unvaccinated and vaccinated children). However, the estimate of overall seropositivity, based on spike antibody positivity, is higher than for the whole population. This is because a higher proportion of children in the study were vaccinated compared to the general population, and overall seropositivity is considerably higher in vaccinated children (35). Thus, our findings highlight the importance of vaccination, in addition to acquired infection, to achieve high levels of seropositivity in children.

This study was able to distinguish children who were seropositive on the basis of SARS-CoV-2 vaccination only, infection only or both vaccination and infection, through measurement of both nucleocapsid antibodies acquired post-SARS-CoV-2 infection and spike antibodies from COVID-19 infection and/or immunization (36). While cumulative nucleocapsid IgG positivity (indicating SARS-CoV-2 infection) remained low at Visit 3 in the autumn of 2021, spike antibody positivity was much higher by then, reflecting rapid uptake of vaccine in the eligible study population at that time (35).

SARS-CoV-2 nucleocapsid IgG antibodies were not found in a considerable minority of children in this study even soon after their infection (within 100 days) and the proportion with nucleocapsid antibodies declined to a low level after 150 days, in contrast to spike IgG antibodies, which stayed positive in nearly all children with and without vaccination or reinfection. This is consistent with findings from adult studies describing a decline in antibody response to the nucleocapsid protein over time (5,37). While a small proportion of participants in this study remained nucleocapsid antibody positive more than 750 days post infection, nearly all seroconverted to negative within 200 days post-infection. In contrast, nearly all SARS-CoV-2 infected unvaccinated participants in our cohort still had spike antibodies throughout the study period, similar to residual blood studies that estimated anti-spike antibodies persistence in more than 95% of participants up to 200 days post infection (10).

Strengths of our study include the longitudinal study design with high participant retention and multiple measurements over multiple waves of the COVID-19 pandemic and the introduction of vaccines, all stratified by age and COVID-19 infection and/or vaccination status. There are multiple reports of point estimates of seropositivity against SARS-CoV-2 in different populations (38,39). Taken together, these show a consistent picture of a gradual rise in population seropositivity during 2020, followed by a sharp rise in 2021 as vaccines were introduced and then levelling off at seropositivity rates nearing 100% in late 2021 into early 2022 after the first Omicron variant wave. However, there are very few published longitudinal studies of repeated SARS-CoV-2 serological measure in children over time, which also include data on incidence of infections and vaccine uptake. Most such studies were conducted before vaccines were introduced (12-18), followed children for less than one year (12-15,18) and were conducted before the Omicron wave of the pandemic (12-18). While serial point prevalence surveys using residual blood sampling have been able to document the rise in seropositivity over the course of the pandemic and were used to inform public health interventions, the clinical and demographic data from the study populations was limited and cannot readily distinguish between seropositivity from infection, vaccination, or both (3,12,40,41).

Limitations of this study include the convenience sampling method of recruitment, as those who are willing to participate in research may be more likely to follow preventative health measures (42), and therefore may have been less likely to be infected with SARS-CoV-2. The sample size was smaller than some comparable serology studies (13,40), most children enrolled in the study were healthy with few underlying conditions, and the initially uninfected study group was not as ethnically diverse as the group who had COVID-19 infections prior to enrollment.

Our results highlight the importance of seropositivity studies as essential tools for monitoring the COVID-19 pandemic and measuring SARS-CoV-2 seropositivity levels (37,43). The pandemic has evolved significantly including the emergence of multiple variant strains of SARS-CoV-2 and the introduction of COVID-19 vaccines. The reduction in diagnostic PCR testing for infection in our region and elsewhere may have contributed to an underestimation of SARS-CoV-2 infections, highlighting the need for serologic testing to accurately measure proportions infected. Although it is not yet clear what a high level of seropositivity means in terms of individual protection against infection and/or severe disease outcomes, some policy makers and health officials now consider that children are largely protected against COVID-19. This is arguable, given that based on our data, a considerable proportion of children in some settings have not had COVID-19 infections, and that uptake of vaccine in young children is lower than for older pediatric groups, and emerging variants continue to contribute towards reinfections. Future longitudinal studies will provide essential information on the incidence and prevalence of SARS-CoV-2 seropositivity and the durability of antibody responses after infection and/or vaccination.

## Data Availability

All data produced in the present study are available upon reasonable request to the authors.

## Acknowledgments

The authors thank all children and families who participated in the study and acknowledge LeeAnn Turnbull and the research staff at Alberta Precision Laboratories-Public Health Laboratory for completing serology testing. We acknowledge contributions by the ACHIEVE Research Team in sample collection (Anna Maria Ang-Becker, Isabelle Banks, Kyu Hwa Lim, Nathalie Uy, and Shairoz Lallany) and participant recruitment and study facilitation (Albert Michael, Candice Jay, Charisse Dominski, Conné Lategan, Julie-Anne Lemay, Nicole MacMillan, Sardar (Shahzeb) Khan, Shannon Pyra, Taylor Cave, and Vivian Ly). We acknowledge non-author members of the Alberta Childhood COVID-19 Cohort Study Team for conceptualization of the seroepidemiology study: Susanne Benseler, Byron Berenger, Francois Bernier (AB3C Study Co-Principal Investigator), Cora Constantinescu, Marvin Fritzler, Otto Vanderkooi.

